# ChatGPT Decision Support System: Utility in Creating Public Policy for Concussion/Repetitive Brain Trauma Associated With Neurodegenerative Diseases

**DOI:** 10.1101/2023.04.17.23288681

**Authors:** Don Comrie

**Author notes:** Corresponding Author: Don Comrie, Neurolabs, Inc., 15 Maiden Lane, Suite 205, New York, NY 10038, USA.

## Abstract

This article evaluates the ChatGPT decision support system’s utility for creating policies related to concussion and repetitive brain trauma associated with neurodegenerative disease risk. It is generally stable and fast. prompt/response pairs (n=259) were examined returning: six prompt response pairs that regenerated (2.31%); one Incorrect Answer; (.38%) one fragment (.38%). Its accuracy, validity, opacity, informational latency and vulnerability to manipulation limits its utility. ChatGPT’s data can be both out-of-date and incomplete which limits its utility use to subject matter experts analyzing expert statements. ChatGPT’s performance is affected by prompts involving stakeholder bias and litigation management, such as race. Nonetheless, ChatGPT demonstrated its ability to respond in both American and British/Australian English with ease. Overall, this study suggests that ChatGPT has limitations that need to be addressed before it can be widely used in decision-making related to concussion and repetitive brain trauma policies.

## Introduction

ChatGPT-3.5 (Generative Pretrained Transformer 3) is a large language model (LLM) AI processing model developed by OpenAI (https://openai.com). The models are capable of generating human-like text that is tailored to appear as a natural conversion. The model is trained on an enormous volume of data. It uses a combination of natural language processing (NLP), machine learning and statistical analysis to interpret prompts, identify relevant information and predict an appropriate response. It is suited to a wide range of applications. Its deployment has generated much consternation. It has not only thrown academic^1^ and scientific research^2^ into turmoil, but also disrupts student assessment.^3^ On the other hand, medical disciplines such as radiology^4^ and urology are evaluating LLMs use. A Urology professor suggests it is ready for application in routinized low-risk tasks in the clinical setting.^5^ In fact, in Australia, University of Sydney medical school incorporates the use of ChatGPT in its courses.^6^

Given medicine’s cautionary interest in ChatGPT and similar systems, it seemed worthwhile to design an experiment to test the utility based on a human evaluation of transcripts of the Australia Senate^7^ Community Affairs References Committee hearing on Concussions and Repeated Head Trauma in Contact Sports. Concussions and repetitive brain trauma have been linked to an increased risk of neurodegenerative diseases such as Chronic Traumatic Encephalopathy (CTE). As a result, policymakers are faced with the challenging task of developing effective policies to mitigate this risk and protect the health of athletes and other individuals who may be at risk of brain injury. It is an area of considerable controversy that is overwhelmed by well-funded experts offering industry friendly responses to public questions.

In that light, the evaluation is intended to assess the utility of ChatGPT to help non-subject experts in legislature and educational settings to develop medical policies. Owing to ChatGPTs access to large volumes of data related to concussion and brain trauma, including medical research, legal cases, and other relevant information, it may become a powerful tool for assessing the potential impact of different policy approaches and evaluating their effectiveness in reducing the risk of neurodegenerative disease.

The paper describes the human evaluation of ChatGPT decision support system and effectiveness in supporting policymakers.

## Experimental Design and Evaluation

### Experimental Framework

The study was conducted using ChatGPT-3.5 (Jan 30 Version). The data was captured between February 1, 2023 and February 12, 2023. A sole input for the experiment was using a Microsoft WIN10.1 Home OS, Firefox 64 bit browser Version 109.0.1, with a standardized Spectrum cable connection operating at 77Mbs/11,6 Mbs download/upload speeds. The prompts were sent from NYC via the internet to ChatGPT. The prompts were predominantly in Australian English with the balance made in American English.

#### Data Analysis

The interactions were analyzed using a combination of qualitative and quantitative methods. Qualitative analysis was used to identify patterns and themes in the responses, while quantitative analysis was used for performance of ChatGPT’s platform.

Prompts are plain text; Response is bold; and Author comments in italics.

### Four-Step Process: The evaluation was conducted in four steps

#### Step 1

Test the accuracy and consistency focused on controversial subject prompts. ChatGPT prompts queried about dementia, especially chronic traumatic encephalopathy, arising from collisions that have conflicting information on the internet. The purpose of this step was to evaluate the consistency of ChatGPT’s responses.

Does playing professional gridiron football result in dementia?

**Studies have shown that repeated head trauma experienced by some professional American football players may increase the risk of developing conditions such as chronic traumatic encephalopathy (CTE) and dementia. However, more research is needed to fully understand the relationship between football and cognitive decline, and not all players will experience these outcomes. It’s important to note that while some players may face higher risks, many factors, such as genetics and lifestyle, can also play a role in the development of cognitive conditions**.

*If Australian Rules Football and Rugby was substituted in the prompt American football, it generated the same response*.

Does playing professional gridiron football result in structural brain damage?

**Repeated head trauma experienced by some professional American football players may lead to structural brain damage, as well as other long-term cognitive problems. Studies have shown that repeated head impacts can result in changes to the brain, including shrinkage of certain regions, reductions in white matter, and the accumulation of abnormal protein deposits. However, it’s important to note that not all football players will experience structural brain damage, and that many other factors, such as genetics and lifestyle, can also play a role in the development of brain problems. Additionally, further research is needed to fully understand the relationship between football and structural brain damage**.

*If Aussie Rules Football and Rugby were substituted in the prompt, ChatGPT generated the same response*.

Is chronic traumatic encephalopathy (CTE) a real disease?

**Yes, chronic traumatic encephalopathy (CTE) is a real disease. It is a progressive degenerative brain disease that is thought to be caused by repetitive head trauma such as concussions or sub-concussive blows. The symptoms of CTE typically include problems with thinking, mood, and behavior, and it is diagnosed post-mortem through an examination of brain tissue. However, much is still unknown about CTE, and ongoing research is being conducted to better understand the disease and how it can be diagnosed and treated**.

*Six responses to six prompts were accurate and consistent*.

#### Step 2

Definition of Terms of Reference used by the legislatures. The Terms of Reference were converted to questions as prompts to evaluate ChatGPT’s definition of terms. If ChatGPT was unable to provide an acceptable response to a question, the prompt was eliminated from the evaluation.^8^

Two Examples are below. Example 1, illustrates ChatGPT response suggesting it can answer policy questions. Example 2, ChatGPT’s response not only answered accurately by including facts that correctly suggest repetitive head injury is a misnomer that should be replaced by brain injury.

### Example 1

Concussions and repeated head trauma in contact sports at all levels, for all genders and age groups, with particular reference to the long and short-term support available to players affected by concussion and repeated head trauma.

**Concussions and repeated head trauma in contact sports, such as football, ice hockey, and boxing, can result in short-term and long-term health problems for players. In the short-term, symptoms of concussions may include headache, dizziness, nausea, and confusion. Over the long-term, repeated head trauma can lead to conditions such as chronic traumatic encephalopathy (CTE), a degenerative brain disease that has been linked to depression, memory loss, and other cognitive impairments**.

**To support players affected by concussions and repeated head trauma, various organizations have implemented protocols for identifying and managing concussions. In the National Football League (NFL), for example, players must undergo a standardized concussion evaluation and be cleared by a team doctor and an independent neurological consultant before returning to play. Additionally, the NFL has established a “88 Plan” that provides financial and medical support to former players suffering from cognitive decline**.

**Similar initiatives and protocols have been implemented at the collegiate and youth levels of football, and in other contact sports. It’s important for players, coaches, and parents to be aware of the risks associated with repeated head trauma and to seek proper medical care and support when necessary**.

### Example 2

Concussions and repeated head trauma in contact sports at all levels, for all genders and age groups, with particular reference to the lack of a consistent definition of what constitutes ‘concussion’.

**Concussions and repeated head trauma in contact sports have become a major concern for all levels, genders, and age groups. One of the challenges in addressing this issue is the lack of a consistent definition of what constitutes a “concussion.” A concussion is a type of traumatic brain injury that occurs as a result of a blow to the head or body**.^**9**,**10**,**11**^ **However, the symptoms and effects of a concussion can vary widely, and there is no standard definition or set of criteria that medical professionals and sports organizations can use to diagnose and treat concussions**.

*Nine terms of references were tested. ChatGPT responses were acceptable*.

#### Step 3

**ChatGPT** - **Self Knowledge**. In this step, participants were asked to pose a series of questions to ChatGPT about its own knowledge and capabilities. The purpose of this step was to evaluate ChatGPT’s probable constraints and limitations.

How does ChatGPT interpret differences between prompts written in US English and written Australian English?

**ChatGPT has been trained on a large corpus of text data, including texts in both US and Australian English. It does not have cultural understanding**.

How does ChatGPT establish the value based on the source of information?

**As an AI language model, ChatGPT does not establish the value of information based on its source. Instead, it processes information based on the language patterns and associations it has learned from its training data, and generates responses based on that information. It is up to the user to evaluate the value and reliability of the information based on their own judgment and critical thinking skills**

Why doesn’t ChatGPT identify the source of its responses?

**ChatGPT generates responses based on the patterns and associations it has learned from its training data, without specifically identifying or attributing the source of that information. This is because ChatGPT is designed to be an artificial intelligence language model that can process and generate text based on its training data and contextual understanding, without specific knowledge of the source of the information**.

**Furthermore, ChatGPT is trained on large amounts of text data from a wide range of sources, which makes it difficult to attribute the specific source of each piece of information it processes. It is up to the user to evaluate the quality and reliability of the responses generated by ChatGPT based on their own critical thinking and judgment**.

Can ChatGPT responses be manipulated by industry long-term data generating campaigns to produce scientific articles, conferences, news publications over a long period of time?

**ChatGPT’s responses can potentially be influenced by industry long-term campaigns to produce scientific articles, conferences, news publications, and other types of information over a long period of time, especially if these campaigns produce a large amount of data that is similar in style and content. This is because ChatGPT is trained on large amounts of text data from a wide range of sources, including scientific articles, news publications, and other forms of text**.

**However, it is important to note that ChatGPT is designed to learn patterns and associations from its training data, and not to favor any particular source or type of information**.

**Furthermore, the quality and reliability of the information that ChatGPT generates depends on the quality and reliability of the data it is trained on. Therefore, if industry campaigns produce biased or misleading information, this could potentially influence the responses generated by ChatGPT**.

**It is up to the users of ChatGPT to critically evaluate the quality and reliability of the responses it generates, and to be aware of the potential influence of industry campaigns or other sources of bias. It is also important to seek out diverse and reliable sources of information to avoid relying solely on one type of data**.

Does ChatGPT use individual query history to adjust its responses?

**As an AI language model, I don’t keep track of individual user query history or personal information. Each query is processed independently, and my responses are based solely on the input I receive and the patterns learned from my training data**.

Is ChatGPT designed to avoid any subjects that could generate litigation against OpenAI?

**OpenAI likely designed ChatGPT with the intention of avoiding any features or behaviors that could lead to legal action or controversy. However, as an AI language model, ChatGPT’s responses are generated based on its training data and may not always reflect OpenAI’s explicit intentions**.

Is ChatGPT designed to avoid responses to controversial or politically charged queries?

**OpenAI likely designed ChatGPT to avoid generating responses that could be considered controversial or politically charged. However, as an AI language model, ChatGPT’s responses are generated based on its training data and may not always reflect OpenAI’s explicit intentions. Additionally, what is considered controversial or politically charged can vary based on individual perspectives and cultural contexts**.

## ChatGPT SUPPORTING NOTES

### Note 1

**“It is up to developers and stakeholders to determine the most appropriate methods and frameworks for addressing bias and discrimination in their models, based on their own values, principles, and ethical considerations**.”

### Note 2

**As a language model developed by OpenAI, I do not have medical advisers or researchers guiding me. Instead, I have been trained on a diverse range of text data to generate answers to questions based on that data. This data includes information about various topics, including health and sports, but it is important to note that my answers are not professional medical advice and should not be taken as such. It is always recommended to consult a qualified healthcare professional for any medical concerns**.

*The notes were responses to work conducted unrelated to this study*.

### 8 prompts - 2 Notes

#### Step 4: Transcript Analysis

In this step, the transcripts of the interactions between ChatGPT and the expert witness and governmental panel (14 persons) were analyzed to identify any patterns or inconsistencies in the responses.

259 prompt response pairs. All prompts were submitted verbatim. Due to length, some prompts some were parsed to test speed and quality of the responses.

### High-Utility Prompt/Response Pairs

#### 12. ChatGPT

##### Senator PRATT

*Can you explain why that is, because there’s evidence—very firm evidence, internationally, in other places—to demonstrate that long-term dementia can occur from small, repeated head knocks et cetera that would not qualify as a concussion, although there may have had to be some other concussion actually occur. That’s what I’m trying to unpack*.

**Yes, the relationship between repeated head impacts and the development of long-term neurological conditions such as CTE is complex and not fully understood. It is believed that the accumulation of subconcussive head impacts, which are defined as head impacts that do not result in symptoms of a concussion, over time can contribute to the development of CTE**.

**Research has shown that repeated head impacts, even if they do not result in a clinically diagnosed concussion, can still result in structural changes in the brain and cause cumulative damage over time. This cumulative damage can eventually lead to the symptoms of CTE**.

**It is important to note that while repeated head impacts are considered a risk factor for CTE, it is still not known exactly how many head impacts are needed, or what level of severity is required, to cause the disease. Research in this area is ongoing, and the scientific community continues to work towards a better understanding of the relationship between repeated head impacts and CTE**.

**In conclusion, while a single concussion may not be considered sufficient for the development of CTE, repeated head impacts, including subconcussive impacts, can increase the risk of developing the condition. Policy and research take into consideration the frequency, severity, and duration of head impacts when evaluating the potential risk for CTE**.

*A very broad response to the prompt, but it is largely complete*.

#### 26. ChatGPT

##### Senator PRATT

No, I wasn’t necessarily implying that. It was more that your code had taken steps to support your research, put a surveillance system in and apply it within and look at the specific context of your sport. That is really where I think your leadership could be applied across other codes. I wondered if you might help us translate that into a recommendation or something from the committee.

**Certainly, I would be happy to help. To translate my code into a recommendation for your committee, here are a few steps that could be taken:**

1. **Start by identifying the specific challenges or problems that you want to address within your sport. This could be related to player performance, injury prevention, or fan engagement, among others**.
2. **Conduct a thorough review of the existing data and technology available to support your goals. This includes looking at the various data sources that are available and determining which are most relevant to your specific needs**.
3. **Based on your research and analysis, develop a comprehensive plan for implementing a monitoring and surveillance system within your sport. This plan should include specific objectives, a timeline for implementation, and a budget for the project**.
4. **Work with your committee to identify any potential risks associated with implementing such a system, such as privacy concerns or data security issues. Develop mitigation strategies to address these risks and ensure that the system is secure and complies with all relevant regulations**.
5. **Once the plan has been approved, work with a team of technology experts to implement the system, including the installation of sensors, the development of data analytics tools, and the integration of data from various sources**.
6. **Regularly evaluate the success of the system and make any necessary adjustments to improve its performance. This could include updating the software, adding new sensors, or incorporating new data sources**.
7. **Finally, share the results of your monitoring and surveillance system with your committee and other stakeholders to demonstrate its impact and help drive support for future initiatives**.

***I****t is very suited to explaining expert testimony in lay terms often in greater detail and clarity. ChatGPT can complete statements of experts evading queries*.

#### Item 100 and 101

##### Senator THORPE

Just to lead on from that: what do you say when sporting organisations put an ex-player’s struggles down to, ’They had mental health issues or struggles’, using this as a reason not to compensate these people?

**In short, the argument that a patient’s struggles are simply due to mental health issues rather than a form of dementia such as CTE is not supported by the medical community. A trained neurologist or neuropsychologist is often the best indicator of the presence of a subtype of dementia and can take into account other factors such as depression or substance use disorder when making a diagnosis. Additionally, the decline in cognitive function over time is a key indicator of CTE, as opposed to other forms of dementia where the decline is typically more rapid. It is important to consider the individual case and take into account all relevant factors when making a diagnosis and determining the best course of treatment**

*Ironically, Australian sports leagues are accepting the existence of CTE*^*12*^ *after two decades of denial, yet same organizations continue gaslighting the association of mental illness and brain injury*.

#### 107. ChatGPT

##### Senator THORPE

Just a follow-on: in your view, what needs to happen to get greater consistency on assessment and return-to-play guidelines for professional and amateur sports?

**To achieve greater consistency in assessment and return-to-play guidelines for professional and amateur sports, a multi-faceted approach is needed. This could include:**

1. **Evidence-based guidelines: Developing return-to-play guidelines that are based on the latest scientific evidence and research on head injuries and concussions. This will help ensure that the guidelines are effective in protecting the health and well-being of athletes**.
2. **Consistent protocols: Ensuring that all sports organizations and governing bodies have consistent protocols in place for managing head injuries and concussions, including assessment, diagnosis, and treatment**.
3. **Standardized testing: Implementing standardized tests for assessing head injuries and determining when an athlete is ready to return to play. This will help to ensure that all athletes are evaluated in the same way, regardless of their level of competition**.
4. **Collaboration between sports organizations: Encouraging collaboration between different sports organizations and governing bodies to develop a common approach to managing head injuries and concussions. This will help to ensure that athletes are protected, regardless of the sport they play**.
5. **Educating athletes and coaches: Providing education and resources for athletes and coaches on the dangers of head injuries and concussions, and how to recognize and manage these injuries**.
6. **Ongoing research: Investing in ongoing research to better understand the long-term effects of head injuries and to develop more effective treatments and return-to-play protocols**.

**By implementing these steps, it may be possible to achieve greater consistency in assessment and return-to-play guidelines for professional and amateur sports, and to better protect the health and well-being of athletes**.

*ChatGPT furnishes very basic but useful suggestions to increase consistency of care*.

#### 135. ChatGPT

Dr Cohen: Let’s not forget concussion doesn’t show up on a CT or an MRI; otherwise there’d be something like the checkout at Woolies every footy game, and every player would go through and have a CT on the way out, and they’d say, ’You’ve got it—you can’t play next week.’ It doesn’t show up, because that damage is down there at an early cellular level.

**That’s correct. Concussions are a type of traumatic brain injury that do not typically show up on imaging tests such as CT scans or MRI scans. The diagnosis of a concussion is usually made based on a combination of symptoms, such as headache, dizziness, confusion, and memory loss, as well as a physical examination and a review of the individual’s medical history. While imaging tests may be used to rule out other types of head injuries, they are not typically used to diagnose a concussion. This is one of the reasons why concussions can be difficult to diagnose and manage. The damage from a concussion occurs at the cellular level, and it’s important to take concussions seriously and to manage them appropriately to prevent long-term consequences**.

*ChatGPT validates Dr. Cohen’s statement*.

#### 137. ChatGPT

Dr Cohen: It’s the long-term stuff that causes what we can finally see, because our instruments aren’t acute enough.

**Yes, the long-term effects of concussions can be severe and are only recently being fully understood. The instruments and techniques used to study the brain have improved in recent years, but we are still limited in our ability to fully understand the impact of concussions on the brain and nervous system. The effects of repeated concussions and subconcussive impacts, which are milder head injuries that do not cause symptoms of a concussion, can accumulate over time and contribute to long-term consequences, such as chronic traumatic encephalopathy (CTE), depression, and other neurological conditions. It’s important to continue to research and study the long-term effects of concussions and to develop better methods of prevention, diagnosis, and management**.

*ChatGPT confirms and extends Dr. Cohen expert statement*.

#### 176. ChatGPT

##### Senator THORPE

Thank you for appearing today on this incredibly important issue that we’re dealing with in sport and other places. This is a question to you all. Who are your medical advisers and researchers? Who are you guided by?

**As a language model developed by OpenAI, I do not have medical advisers or researchers guiding me. Instead, I have been trained on a diverse range of text data to generate answers to questions based on that data. This data includes information about various topics, including health and sports, but it is important to note that my answers are not professional medical advice and should not be taken as such. It is always recommended to consult a qualified healthcare professional for any medical concerns**.

*ChatGPT confirms it is not a medical adviser and the user should rely on a qualified healthcare professional*.

#### 212. ChatGPT

CHAIR (Senator Rice) What’s your perspective on the level of focus of the NRL in terms of making sure that the clubs are reporting and telling the clubs that this is a real priority and that they’ve got to resource it properly?

**As an AI language model, I do not have personal opinions; however, I can provide information based on the available data**.

**The National Rugby League (NRL) has a responsibility to ensure the health and safety of its players, and proper documentation and reporting of injuries is a crucial aspect of this. It appears that the NRL has made player health and safety a priority and has taken steps to educate the players and promote the importance of reporting injuries**.

**The level of focus that the NRL places on ensuring that the clubs are reporting injuries properly is likely to vary based on the specific circumstances and resources available. It is possible that the NRL may have implemented policies and procedures to encourage and support clubs in their reporting efforts, but without access to more specific information, it is difficult to say for sure**.

**In any case, it’s important for the NRL to work closely with the clubs and other relevant stakeholders to ensure that proper documentation and reporting of injuries is a priority, and that the necessary resources are available to support this. This can help to promote transparency and accountability, and ensure that the health and safety of the players is protected**

*ChatGPT is a good explainer. It fills in for the evading expert with an expansive policy*.

#### 229. ChatGPT

Mr Busch: We’ve seen this at the highest levels of the sport; this isn’t just a resourcing challenge. We’ve seen it at the World Cup, where we’ve seen players concussed. There have been very quick assessments done. They’ve come back on and realised they’re essentially not in a good state and subsequently have had to come off. To our minds, that speaks to the fact that the laws of the game aren’t facilitating a long enough opportunity to allow for that person to be removed and be assessed for actually going back out, with everybody observing this individual and picking up that they’re in no state to be on that pitch. In that same time, they could potentially have suffered a subsequent concussion, which we know would be far more detrimental to them.

**It’s concerning to hear that players have returned to play despite being suspected of having a concussion, as this can put their health and safety at risk. The fact that these incidents have happened at the highest levels of the sport highlights the importance of making changes to ensure that players receive proper care and evaluation following a head injury**.

**In order to address these issues, it may be necessary to revise the laws of the game and provide more time and resources for on-field evaluations and assessments. This could include the introduction of the substitutive interchange concussive rule that you mentioned earlier, or other measures that would help ensure that players receive proper care and evaluation**.

**Ultimately, the health and safety of athletes must be a top priority, and it’s important to make changes that will help prevent these kinds of incidents from happening in the future. This could involve working with governing bodies, medical professionals, and other stakeholders to improve protocols and procedures for evaluating and treating suspected concussions**.

*ChatGPT takes the expert’s opinion to task*.

#### 231. ChatGPT

Mr Busch: Not all things are equal. The risk of someone doing that versus having their whole life negatively impacted do not have the same weighting.

**Absolutely, player safety should always be the main concern. The consequences of not properly evaluating and treating a player who has suffered a concussion can be long-lasting and potentially life-altering. It’s important that the rules and regulations are in place to minimize these risks and prioritize the health and well-being of the players. The player’s quality of life and future should be taken into consideration over any strategic or competitive advantages that may be gained from not properly evaluating them for a concussion**.

*ChatGPT takes the expert’s opinion to task*.

##### Senator THORPE

Are the same safety mechanisms in place for the women?

#### 241. ChatGPT

*Are the same safety mechanisms in place for the women?*

**The safety mechanisms and protections in place for female athletes in rugby league can vary depending on the league and the country in which they play. However, in many cases, female athletes may not have access to the same level of safety mechanisms and protections as their male counterparts due to unequal pay and less robust insurance policies**.

**It is important to note that the issue of unequal support and protection for female athletes is not limited to rugby league, but is a wider issue in professional sports. It is crucial for sports**

**organizations and player associations to address these inequalities and work towards providing equal support and protection for all athletes, regardless of gender**.

**In recent years, there has been increasing advocacy for equal pay and support for female athletes, and it is hoped that these efforts will lead to improved safety mechanisms and protections for female athletes in rugby league and other sports**.

*ChatGPT gives a fair description of women’s health and safety in Rugby*.

### Three Multi-pair Exchanges Illustrates ChatGPT Deficits

ChatGPT vulnerable to **Argument from authority** (*argumentum ab auctoritate)*

ChaptGPT presents Concussion in Sports Group (CISG) as a standard of traumatic brain injury and neurodegenerative science written by prominent scientists. In fact, currently available public data cast a pall over ChatGPT responses. For example, industry experts and defense lawyers’ experts in brain injury cases cite Concussion In Sports Concussion Consensuses as justification for statements undermining valid science and impeaching alternate options. ChatGPT seems to have not been trained that CISG Consensuses are not peer-reviewed.^13,14,15,16,17^ Questions about the standing of the CISG consensus are not new^18^ nor isolated.^19^ Further, the authors have many commonly known conflicts of interest. By the 5th Consensus (2017), the authors were compelled to report conflicts although most were incomplete disclosures. Finally, the founder and organizer of CISG is a known plagiarist with 10 papers^20^ retracted and over 40 papers^21^ having letters of concern. It was widely covered in the scientific^22^ and the sports press.^23^

ChatGPT is an AI language that model is programmed to provide responses based on the input it receives. It relies on black box algorithms that do not identify the source of the means of constructing responses.

In conclusion, ChatGPT can provide useful insights and information on a variety of topics. It is important to critically evaluate the information presented and consider multiple sources of evidence before forming any conclusions.

### Generalized vs. Specific - Issue of Sports Players Union

Unions have grown in popularity and stature in recent years.^24^ In a series of ChatGPT prompts and responses it suggests unions are good for worker health and well-being. While this may be true to the general labor population, professional collision sports athletes have expressed disappointment with their unions. Further, the issues faced by professional collision sports athletes may require a different approach from that of other industries, and their unions must tailor its actions to these challenges.

Complaints of poor medical care in collisions sports is reported in US, UK, Ireland, Australia,^25^ and New Zealand players for lack of protection from brain injury and related to brain injury. The US NFL Players

Association (NFLPA) has a long history of litigation with its members and retirees. NFLPA is currently being sued in multiple jurisdictions over honoring claims long-term disability benefits.^26, 27^

### More is Less in Large Texts - LLMs have documented issues managing long dialogs^28^

Historically LLMs have documented issues managing long dialogs. Open AI failed to post size limits for prompts. In that light, each prompt was entered verbatim initially. Long prompts extracted from hearings submitted to ChatGPT returned a very short responses. Often ChatGPT responses tend to be not only short but of low quality. It seems to have difficulty formulating coherent responses due to the complexity and variety of the information presented. Even If a long prompt is divided into smaller logical prompts continued to produce a low quality or short results. Clearly, ChatGPT continues to struggle with not only long dialogs and but also poorly constructed statements.

## Discussion

ChatGPT 3.5 is a robust text generator. In the very narrow area of inquiry, ChatGPT could furnish usable responses to both controversial and settled terms. It uses black box algorithms that limit the utility in creating policies. LLM’s have known issues with accuracy^29^, validity^30^,and language.^31^ A more complete assessment of deficits was prepared by Sejnowski.^32^ When prompted, ChatGPT responses confirmed its shortfalls.

The experiment identified issues either not identified or not fully described elsewhere. Susceptible to Argument from authority (*argumentum ab auctoritate*) applies to many problems. A means of dealing with the problem is necessary but it requires a depth of knowledge to resolve. The problem of specific/general paradox again requires in depth knowledge. The first two issues might be addressed by applying chain of thought prompting.^33^ Large scale text analysis is currently beyond LLM’s capability at the present time.

The evaluation discovered ChatGPT could furnish suitable responses to prompts evaded by industry representatives. It wasn’t entirely surprising since it can respond based on vast data that trained it. On the other hand, it was surprising that ChatGPT responses to industrial prompts compromised of baseless, unsubstantiated, unsupported assertions provoked a dress down from the LLM. It is unclear what generated the result, but it suggests a layer of coding yet to be disclosed to the public.

It should be noted that text analysis using input for multiple parties (14 parties) was accomplished. It did not induce hallucinations, but ChatGPT mirroring (response emulates the prompt input), did expose the evaluator to textual schizophrenia when reviewing the ouput. Finally, Australian Senators not only maintain high levels of decorum but also makes statements laced with wit, humor and gentle mockery. ChatGPT failed to neither comprehend pithy prompts nor embrace the nuances.

## Limitations

First, the study was limited to narrow medical policies regarding traumatic brain injury and neurodegenerative disease, and therefore the results may not be applicable to other medical policies or conditions. Second, the study primarily focused on a cohort of adult males employed or retired from professional collision sports, which may limit the generalizability of the findings to other populations, such as women or non-athletes.

Furthermore, the study relied on prompts furnished by policymakers who were well-educated about the topic, which may have influenced the responses of the expert witnesses. Additionally, the fact the study utilized sworn testimony from these experts, many of whom were well-compensated, may have also introduced bias or lowered the quality of the prompts analyzed.

Finally, while the experiment demonstrated high levels of narrow utility tailored to a specific audience, the generalizability of the results to other populations, subjects, topics or contexts remains uncertain.

On the other hand, similar studies may be afflicted by the deficiencies identified in the experiment. Future research should aim to replicate these findings using similar analyses while examining a broader range of populations and medical issues and controlling the issues that degraded the results.

## Conclusion

ChatGPT is a communications accomplishment. Its fluid text generation of compressible responses to prompts required nearly 45 years to perfect beam searches.^34^ ChatpGPT is a real achievement but one should acknowledge machine intelligence was first broached in public by Alan Turing in 1947.^35^ Artificial Intelligence was coined as part of the proposal of the Dartmouth Summer Research Project on Artificial Intelligence in 1955.^36^ The 75 years of progress has been stunted. It has been marked by fits and starts while lacking a transformational event. Today, ChatGPT’s rapid adoption by the public has fueled demands for a uniform adoption of principles^37^ to regulate AIs growth. A petition has been distributed among technologists to suggest AI research and development be paused due to a lack of understanding its effects.^38^ On the other hand, AI has suffered from an overestimation of its performance projecting mass labor market disruption^39^ while failing to realize the difficulty to accomplish human replacement. The experiment conducted used transcripts from a governmental hearing. Using ChatGPT decision support system for concussion/repetitive brain trauma associated with neurodegenerative disease risk policies is premature. First, since ChatGPT relies on black box algorithms, its application to medical policy should be pursued with caution.^40^ ChatGPT deficits related to accuracy, validity, opacity, bias, manipulation, completeness, and latency were evident during the evaluation. ChatGPT’s current greatest utility is in confirming an ethical subject matter expert’s views or impeaching so-called experts that evade, dissemble, or mislead on established settled facts. Nonetheless, all responses should be reviewed and validated by a subject matter expert. Experts should follow Pascal^41^ and take their time preparing a prompt. ChatGPT prompts should be clear, concise and focused on a specific topic to obtain best results.

Finally, Artificial Intelligence remains in a gestational phase. ChatGPT is stable, fast and robust. Its text generation capability is extraordinary. Numerous problems will be addressed in upcoming years. Nonetheless, ChatGPT will always be limited to what contained iinits training dataset. Developers are already classifying ChatGPT as a communications interface to bridge the world between humans and computational platforms like Wolfram’s Alpha 1.^42^ One must acknowledge the limitations of AI. Picasso once said, “What good are computers? They can only give you answers.”^43^ Until AI systems can form important questions, humans shouldn’t panic. In fact once a machine is capable of producing important questions it is unclear whether humans should panic.

## Data Availability

No

## Patient consent

Not applicable.

## Data Availability Statement

No data are available.

## References

1 Thorp, H. H. (2023). ChatGPT is fun, but not an author. Science, 379(6630), 313–313.

2 Nature Editorial. (2023). Tools such as ChatGPT threaten transparent science; here are our ground rules for their use. Nature, 613(7945), 612–612.

3 Rudolph, J., Tan, S., & Tan, S. (2023). ChatGPT: Bullshit spewer or the end of traditional assessments in higher education?. Journal of Applied Learning and Teaching, 6(1).

4 Kitamura, F. C. (2023). ChatGPT is shaping the future of medical writing but still requires human judgment. Radiology, 230171.

5 Gabrielson, A. T., Odisho, A. Y., & Canes, D. (2023). Harnessing Generative Artificial Intelligence to Improve Efficiency Among Urologists: Welcome ChatGPT. The Journal of Urology, 10–1097.

6 Harris, C. (2023, March 14). Medical science students were told to use CHATGPT. this is what it wrote. The Sydney Morning Herald. Retrieved March 20, 2023, from https://www.smh.com.au/national/nsw/medical-science-students-were-told-to-use-chatgpt-this-is-what-it-wrote-20230309-p5cqs4.html

7 Parliament of Australia. “Community Affairs References Committee - Concussions and Repeated Head Trauma in Contact Sports.”Aph.gov.au, Parliament of Australia, 30 Jan. 2023, Accessed 24 Mar. 2023.

8 Commonwealth Parliament; “Terms of Reference.” Parliament of Australia, Commonwealth Parliament; Parliament House, Canberra, ACT, 2600; Contact=+61 2 6277 7111, 1 Dec. 2022, https://www.aph.gov.au/Parliamentary_Business/Committees/Senate/Community_Affairs/Headtraumainsport/Terms_of_Reference.

9 Tator, C. H. (2013). Concussions and their consequences: current diagnosis, management and prevention. Cmaj, 185(11), 975–979.

10 Hynes LM, Dickey JP. (2006). Is there a relationship between whiplash-associated disorders and concussion in hockey? A preliminary study. Brain Inj 006;20:179–88

11 Cernak, I., & Noble-Haeusslein, L. J. (2010). Traumatic brain injury: an overview of pathobiology with emphasis on military populations. Journal of Cerebral Blood Flow & Metabolism, 30(2), 255–266.

12 Convery, Stephanie. “NRL and Football Australia Accept Link between Head Trauma and CTE.”The Guardian, Guardian News and Media, 1 Mar. 2023, https://www.theguardian.com/sport/2023/mar/01/nrl-and-football-australia-accept-link-between-head-trauma-and-cte.

13 Aubry, M., Cantu, R., Dvorak, J., Graf-Baumann, T., Johnston, K., Kelly, J., … & Schamasch, P. (2002). Summary and agreement statement of the first International Conference on Concussion in Sport, Vienna 2001. The Physician and sportsmedicine, 30(2), 57–63.

14 McCrory, P., Johnston, K., Meeuwisse, W., Aubry, M., Cantu, R., Dvorak, J., … & Schamasch, P. (2005). Summary and agreement statement of the 2nd International Conference on Concussion in Sport, Prague 2004. British journal of sports medicine, 39(4), 196–204.

15 McCrory, P., Meeuwisse, W., Johnston, K., Dvorak, J., Aubry, M., Molloy, M., & Cantu, R. (2009). Consensus statement on Concussion in Sport–the 3rd International Conference on Concussion in Sport held in Zurich, November 2008. South African Journal of sports medicine, 21(2).

16 McCrory, P., Meeuwisse, W. H., Aubry, M., Cantu, R. C., Dvořák, J., Echemendia, R. J., … & Turner, M. (2013). Consensus statement on concussion in sport: the 4th International Conference on Concussion in Sport, Zurich, November 2012. Journal of athletic training, 48(4), 554–575.

17 McCrory, P., Meeuwisse, W., Dvorak, J., Aubry, M., Bailes, J., Broglio, S., … & Vos, P. E. (2017). Consensus statement on concussion in sport—the 5th international conference on concussion in sport held in Berlin, October 2016. British journal of sports medicine, 51(11), 838–847.

18 Casper, S. T., Bachynski, K. E., Buckland, M. E., Comrie, D., Gandy, S., Gates, J., … & Finkel, A. M. (2021). Toward complete, candid, and unbiased international consensus statements on concussion in sport. Journal of Law, Medicine & Ethics, 49(3), 372–377

19 McNamee, M., Anderson, L. C., Borry, P., Camporesi, S., Derman, W., Holm, S., … & Weed, M. (2023). Sport-related concussion research agenda beyond medical science: culture, ethics, science, policy. Journal of medical ethics.

20 Macdonald, H., Ragavooloo, S., Abbasi, K., & Drezner, J. A. (2022). Update on the investigation into the publication record of former BJSM editor-in-chief Paul McCrory. British Journal of Sports Medicine, 56(23), 1327–1328.

21 Mahase, E. (2022). British Journal of Sports Medicine retracts editorial by former editor over plagiarism concerns.

22 Kincaid, Ellie. “Concussion Researcher Paul McCrory Earns Nine More Retractions, Nearly 40 Expressions of Concern.”Retraction Watch, 10 Oct. 2022, https://retractionwatch.com/2022/10/10/concussion-researcher-paul-mccrory-earns-nine-more-retractions-nearly-40-expressions-of-concern/.

23 Bull, Andy. “Latest Mccrory Retractions Leave Sport Facing a Reckoning over Concussion | Andy Bull.”The Guardian, Guardian News and Media, 13 Oct. 2022, https://www.theguardian.com/sport/blog/2022/oct/13/latest-mccrory-retractions-leave-sport-facing-a-reckoning-over-concussion.

24 McCarthy, Justin. “U.S. Approval of Labor Unions at Highest Point since 1965.”Gallup.com, Gallup, 25 Jan. 2023, https://news.gallup.com/poll/398303/approval-labor-unions-highest-point-1965.aspx.

25 Wilson, C. (2023, April 14). 14 West Coast players take out trauma insurance to tackle concussion threat. The Age. https://www.theage.com.au/sport/afl/14-west-coast-players-take-out-trauma-insurance-to-tackle-concussion-threat-20230413-p5d07p.htm

26 Cloud v. The Bert Bell/Pete Rozelle NFL Player Ret. Plan, Civil Action 3:20-CV-1277-S (N.D. Tex. Jun. 21, 2022)

27 Alford v. NFL Player Disability & Survivor Benefit Plan, Civil action No. 1:23-cv-00358 (D. Md., 2/9/23).

28 Valvoda, J., Fang, Y., & Vandyke, D. (2022). Prompting for a conversation: How to control a dialog model?. arXiv preprint arXiv:2209.11068.

29 Bang, Y., Cahyawijaya, S., Lee, N., Dai, W., Su, D., Wilie, B., … & Fung, P. (2023). A multitask, multilingual, multimodal evaluation of chatgpt on reasoning, hallucination, and interactivity. arXiv preprint arXiv:2302.04023 30

30 Peng, B., Galley, M., He, P., Cheng, H., Xie, Y., Hu, Y., … & Gao, J. (2023). Check your facts and try again: Improving large language models with external knowledge and automated feedback. arXiv preprint arXiv:2302.12813.

31 Weidinger, L., Mellor, J., Rauh, M., Griffin, C., Uesato, J., Huang, P. S., … & Gabriel, I. (2021). Ethical and social risks of harm from language models. arXiv preprint arXiv:2112.04359.

32 Sejnowski, T. J. (2023). Large language models and the reverse turing test. Neural Computation, 35(3), 309–342

33 Wei, J., Wang, X., Schuurmans, D., Bosma, M., Chi, E., Le, Q., & Zhou, D. (2022). Chain of thought prompting elicits reasoning in large language models. arXiv preprint arXiv:2201.11903.

34 Goodman, G., & Reddy, R. (1977). Alternative control structures for speech understanding systems.

35 Turing, A. M. (1995). Lecture to the London Mathematical Society on 20 February 1947. MD COMPUTING, 12, 390–39

36 McCarthy, J., Minsky, M. L., Rochester, N., & Shannon, C. E. (2006). A Proposal for the Dartmouth Summer Research Project on Artificial Intelligence, august 31, 1955. AI magazine, 27(4), 12–12.

37 “AI Principles.” Future of Life Institute, 22 Mar. 2023, https://futureoflife.org/open-letter/ai-principles/.

38 “Pause Giant AI Experiments: An Open Letter.” Future of Life Institute, Future of Life Institute, 13 Apr. 2023, https://futureoflife.org/open-letter/pause-giant-ai-experiments/.

39 Eloundou, T., Manning, S., Mishkin, P., & Rock, D. (2023). Gpts are gpts: An early look at the labor market impact potential of large language models. arXiv preprint arXiv:2303.10130.

40 Durán, J. M., & Jongsma, K. R. (2021). Who is afraid of black box algorithms? On the epistemological and ethical basis of trust in medical AI. Journal of Medical Ethics, 47(5), 329–335.

41 quoteresearch, Author. “If I Had More Time, I Would Have Written a Shorter Letter.” Quote Investigator, 19 July 2022, https://quoteinvestigator.com/2012/04/28/shorter-letter/.

42 Stephen Wolfram on AI’s rapid progress & the “Post-Knowledge Work Era” | E1711-https://www.youtube.com/watch?v=F5tXWmCJ_wo

43 Fiffield, William. “William Fifield Interviews Pablo Picasso.”Paris Review, 1964.

